# Robotic RNA extraction for SARS-CoV-2 surveillance using saliva samples

**DOI:** 10.1101/2021.01.10.21249151

**Authors:** Jennifer R. Hamilton, Elizabeth C. Stahl, Connor A. Tsuchida, Enrique Lin-Shiao, C. Kimberly Tsui, Kathleen Pestal, Holly K. Gildea, Lea B. Witkowsky, Erica A. Moehle, Shana L. McDevitt, Matthew McElroy, Amanda Keller, Iman Sylvain, Ariana Hirsh, Alison Ciling, Alexander J. Ehrenberg, IGI SARS-CoV-2 consortium, Bradley R. Ringeisen, Garth Huberty, Fyodor D. Urnov, Petros Giannikopoulos, Jennifer A. Doudna

## Abstract

Saliva is an attractive specimen type for asymptomatic surveillance of COVID-19 in large populations due to its ease of collection and its demonstrated utility for detecting RNA from SARS-CoV-2. Multiple saliva-based viral detection protocols use a direct-to-RT-qPCR approach that eliminates nucleic acid extraction but can reduce viral RNA detection sensitivity. To improve test sensitivity while maintaining speed, we developed a robotic nucleic acid extraction method for detecting SARS-CoV-2 RNA in saliva samples with high throughput. Using this assay, the Free Asymptomatic Saliva Testing (IGI-FAST) research study on the UC Berkeley campus conducted 11,971 tests on supervised self-collected saliva samples and identified rare positive specimens containing SARS-CoV-2 RNA during a time of low infection prevalence. In an attempt to increase testing capacity, we further adapted our robotic extraction assay to process pooled saliva samples. We also benchmarked our assay against the gold standard, nasopharyngeal swab specimens. Finally, we designed and validated a RT-qPCR test suitable for saliva self-collection. These results establish a robotic extraction-based procedure for rapid PCR-based saliva testing that is suitable for samples from both symptomatic and asymptomatic individuals.

## Introduction

The COVID-19 pandemic has motivated extensive scientific efforts to develop vaccines, therapeutics and diagnostics. Detection of asymptomatic individuals carrying the virus is essential for informing public health interventions and minimizing the spread of SARS-CoV-2. Universities seeking to maintain a level of in-person activity have sought to implement asymptomatic screening methods that can help guard against asymptomatic spread in a population that is more likely than the general population to be asymptomatic or exhibit mild symptoms when infected^1–3^. Nasal swab or saliva samples obtained by health practitioners are time-consuming and expensive to collect, motivating our clinical testing laboratory and many others to explore the utility of self-collected specimens^4,5^. Saliva self-collection enabled increased population sampling capacity but, in turn, increased the demand for a rapid yet sensitive PCR-based testing protocol. To address these challenges, we developed an automated procedure for extraction and detection of SARS-CoV-2 RNA in saliva as part of the Innovative Genomics Institute/UC Berkeley Free Asymptomatic Saliva Testing (IGI-FAST) research study^6,7^. Here we describe a robust, high-throughput saliva testing strategy that includes viral inactivation requirements for handling potentially infectious samples, a fully automated RNA-extraction-based detection method, and attempts at saliva sample pooling. This procedure, which is suitable for at-home collection, enabled surveillance testing of 11,971 samples from asymptomatic individuals and identification of SARS-Cov-2-positive specimens even when the prevalence of viral infection was <1% in the East Bay community^8^.

## Results

### Extraction-based detection of viral RNA in saliva

We set out to establish a robotic RNA extraction pipeline to detect SARS-CoV-2 RNA in saliva specimens by adapting our previously described assay for detecting SARS-CoV-2 RNA from healthcare provider-collected respiratory swabs^6^ (**Fig. 1a**). We selected DNA Genotek OMNIgene collection tubes (OM-505) as they had previously been approved for detection of SARS-CoV-2 from saliva^9^. To ensure a maximally safe environment for laboratory staff and volunteers and to comply with UC Berkeley Environmental Health & Safety (EH&S)requirements, we created a protocol that would prevent infectious SARS-CoV-2 in collected samples from entering the diagnostic laboratory. To test the capacity of OM-505 collection tubes to completely inhibit viral infectivity, tissue culture supernatant containing SARS-CoV-2 was combined with OMNIgene sample stabilization fluid at a 1:1 ratio, incubated either at room temperature (22°C) for 60 minutes or at 65°C for 30 minutes, and then tested whether infectious virus remained following treatment using a cytopathic effect (CPE) assay. The sample incubated at 22°C but not at 65°C induced cell death in inoculated Vero-E6 cells, indicating a 65°C incubation was required to eliminate infectious virus (**Fig. 1b**). As DNA Genotek recommends pre-incubation of samples at 50°C for 120 minutes to activate embedded collection tube proteases, this resulted in a two-step pre-extraction sample incubation: 50°C for 120 minutes followed by 65°C for 30 minutes. While all experiments presented in this work use the two-step pre-extraction sample incubation, we have since repeated the viral inactivation study and have found that 120 minutes at 50°C alone is sufficient for complete viral inactivation (**Supplemental Figure 1**).

**Figure 1:**
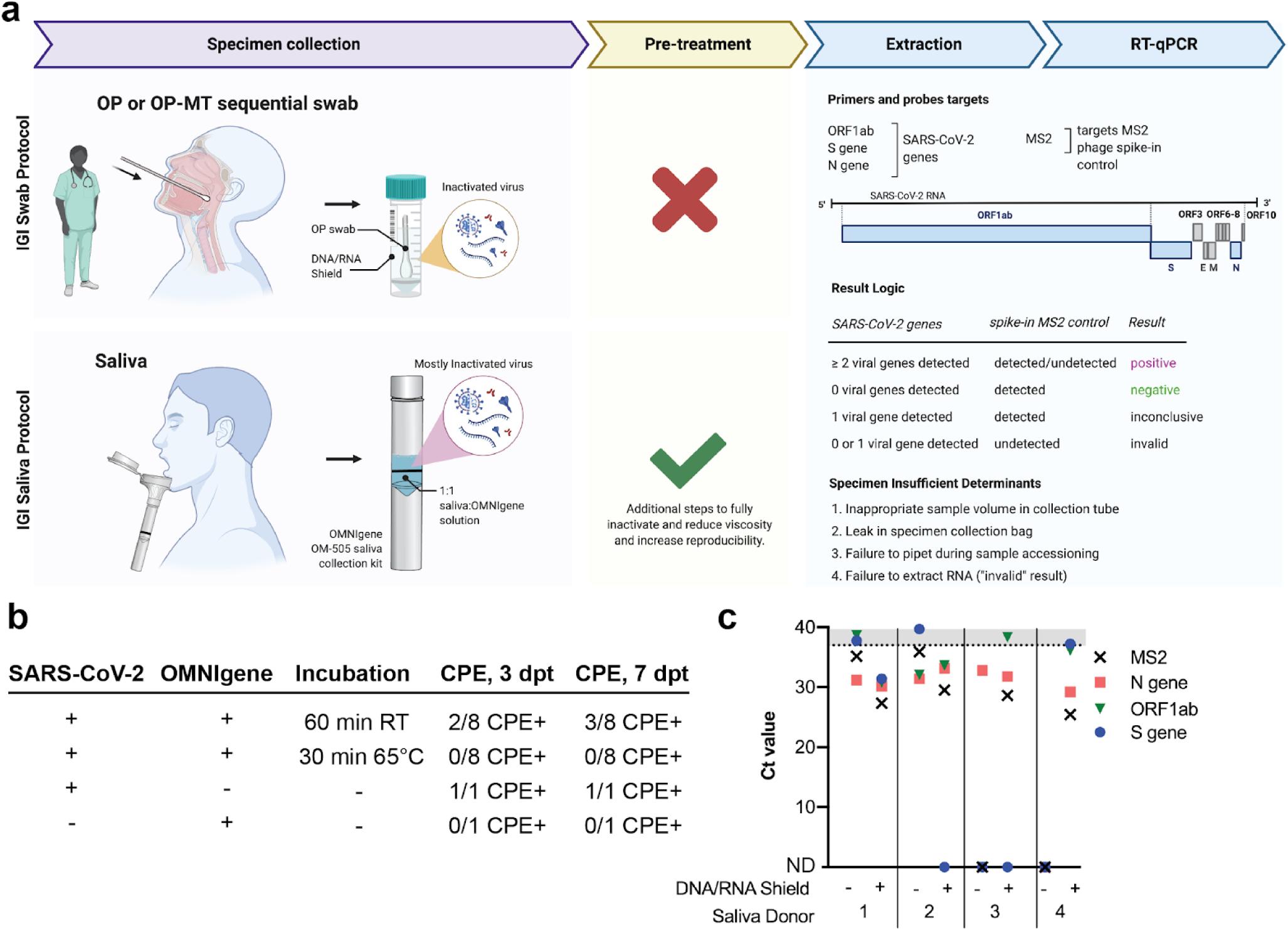
Overview of nucleic acid extraction from saliva specimens. **a**, Overview of Innovative Genomics Institute’s (IGI’s) specimen processing pipeline for both swab and saliva samples. OP = oropharyngeal. OP-MT = oropharyngeal-mid turbinate. **b**, Cultured SARS-CoV-2 (1.58×10^6^ TCID50/ml) was mixed 1:1 with OMNIgene solution present in OM-505 collection tubes to test incubation conditions that inactivate viral replication. Samples were either held at room temperature (RT) or incubated at 65°C for the indicated length of time before being applied to Vero-E6 cells. Cytopathic effect (CPE) was quantified at 3 and 7 days post treatment (dpt). **c**, 3:2 dilution of saliva samples with DNA/RNA Shield improves detection of spiked-in SARS-CoV-2 RNA or MS2 in four saliva donors.

We next developed procedures for robotic sample plating and RNA extraction from saliva samples. Processing saliva:OMNIgene samples (hereafter referred to simply as “saliva samples”) directly through our validated swab-based automated sample plating, extraction and RT-qPCR protocol^6^ was initially unsuccessful. The sample-to-sample variability in saliva viscosity caused two significant impediments to the assay. First, variability in sample viscosity inhibited accurate robotic pipetting during sample plating, resulting in an unusable specimen and a consequent “specimen-insufficient” result. Second, of those samples that were able to be arrayed, we observed that sample viscosity further reduced the efficacy of the collection of nucleic acid-bound beads during magnetic concentration, resulting in loss of bead-bound sample during extraction. The ThermoFisher TaqPath COVID-19 assay relies upon detection of a spiked-in MS2 internal control to quantify the efficiency of RNA extraction, and initial experiments with saliva samples resulted in poor MS2 detection, consistent with our observation of bead loss (**Fig. 1c**, DNA/RNA Shield (-) samples).

Motivated by the success of our nasal swab-based assay with specimens collected in a chaotropic solution (DNA/RNA Shield, Zymo Research), we hypothesized that saliva specimen performance could be improved through dilution with DNA/RNA Shield without compromising sensitivity. Counterintuitively, but in accordance with our hypothesis, we found that diluting saliva samples with DNA/RNA Shield improved RNA detection, with most spiked-in viral RNA and MS2 internal controls being detected at a lower cycle threshold (Ct) or detectable when previously undetectable (**Fig. 1c**). By performing a titration curve mixing saliva with either DNA/RNA Shield or phosphate buffered saline (PBS), we found that a 3:1 ratio of DNA/RNA Shield:saliva sample resulted in optimal detection of spiked-in MS2 following extraction (**Supplemental Figure 2**). As specimen dilution decreased the total amount of saliva transferred for extraction, we subsequently doubled both the sample input volume and the volume of MagMax DNA/RNA-binding beads in our robotic nucleic acid extraction. The full protocol for robotic nucleic acid extraction from saliva is outlined in **Supplemental Figure 3**.

Having established a protocol compatible with both viral inactivation and robotic sample processing, we next evaluated the robustness of the IGI SARS-CoV-2 saliva assay for contrived virus-positive saliva specimens. We obtained saliva from four donors who had recently tested negative in an oropharyngeal swab qPCR-based test. The donated saliva was mixed 1:1 in OM-505 collection tubes to mimic a specimen collected according to the OMNIgene kit collection instructions. To determine the limit of detection of our assay, we generated titration curves by adding known quantities of either Thermo Fisher TaqPath SARS-CoV-2 positive control RNA or heat-inactivated SARS-CoV-2 virus to the prepared donor clinical matrix. The limit of detection (LoD, concentration at which all biological replicates would be called positive by detection of two or more viral genes with a Ct<37, see **Fig. 1a**) for this assay was determined to be 3×10^3^ RNA copies/mL (**Fig. 2a**) and 5 TCID50/mL (fifty-percent tissue culture infective dose) (**Fig. 2b**). We confirmed the robustness of our assay by testing 20 saliva specimens to which we added either viral RNA or heat-inactivated virus at 2x and 5x LoD into saliva collected from donors who had recently received negative results using respiratory swab-based tests (**Fig. 2c**). All contrived saliva specimens were experimentally determined to be positive, indicating that this assay appropriately detects the presence of SARS-CoV-2 from RNA robotically extracted from saliva. We further tested the reproducibility of our assay by adding 3×10^3^ RNA copies/mL SARS-CoV-2 RNA (1x LoD) to 20 negative saliva specimens. At the determined LoD, 19/20 contrived positive samples were called positive (95%) and 1/20 called invalid due to failure to extract MS2 (**Fig. 2d**). In an intermediate precision assay, this experiment was repeated with the same samples on a different day and returned the same results, suggesting that the dropout of sample 10 was due to factors inherent to the sample itself and was not due the assay (data not shown). Together, we successfully adapted the IGI’s swab-based robotic RNA extraction pipeline for the sensitive detection of SARS-CoV-2 from saliva and achieved an LoD that compares favorably to commercially-available diagnostic assays (**Fig. 5e**).

**Figure 2:**
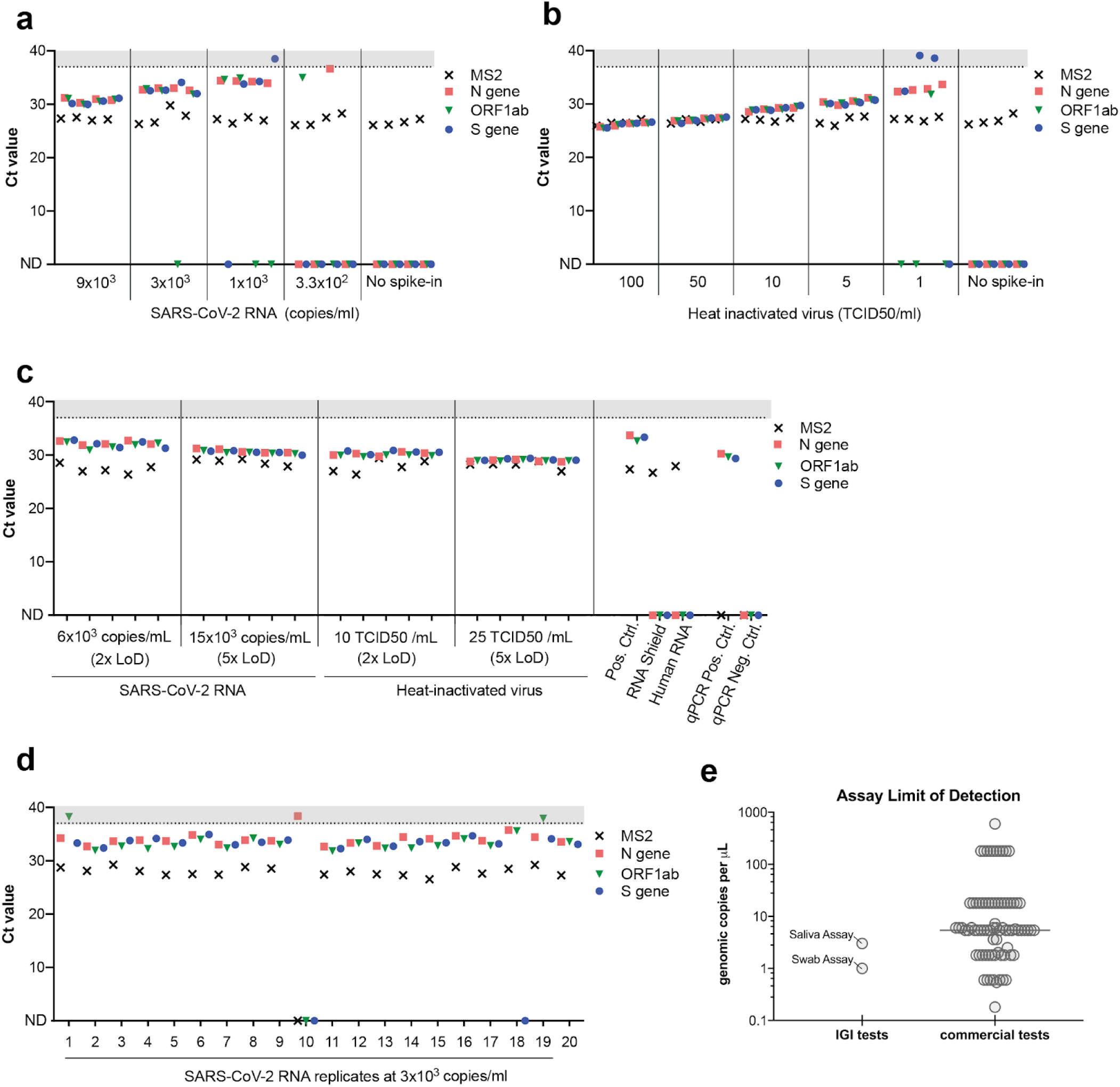
Validation of the IGI’s assay for detecting SARS-CoV-2 RNA from saliva. Saliva collected from four unique donors (negative for SARS-CoV-2 by swab) was used to generate a titration curve of ThermoFisher COVID-19 Positive Control SARS-CoV-2 RNA (**a**) or heat-inactivated virus (**b**) to determine the assay’s limit of detection (LoD). **c**, SARS-CoV-2 RNA or heat-inactivated virus was spiked in at 2x or 5x the LoD into 20 unique saliva samples previously determined to be negative for SARS-CoV-2. **d**, SARS-CoV-2 RNA was spiked into 20 unique saliva samples previously determined to be negative for SARS-CoV-2 at 1x LoD (3×10^3^ copies/ml). A positive extraction control (Pos. Ctrl) negative extraction controls (DNA/RNA Shield, Human RNA) and qPCR controls all returned expected results (**c**). Ct values >37 are shaded in gray. Undetected Ct values are plotted as zero and designated by “ND”, not detected. **e**, Limit of detection (RNA copies/µl) comparison of commercial assays and the IGI saliva and IGI swab tests generated from the FDA website^10^.

### Results from the IGI-FAST research study

After developing an initial procedure that performed robustly in the laboratory, we implemented our approach in a real-world setting through an institutional review board-supervised research study. This study sought to both validate our procedures, bring free, asymptomatic testing to our campus, and develop a streamlined collection operation. Implementation of the initial protocol — which lacked the 50°C pre-incubation and sample dilution with DNA/RNA Shield — returned a high number of “Specimen Insufficient” samples in our IGI-FAST cohort (**Fig. 2a**, weeks 1-3). A “Specimen Insufficient” outcome can result from either (1) an inappropriate sample volume in the collection tube; (2) a leak in the specimen collection bag; (3) a failure to pipet during sample plating; or (4) a failure to extract RNA (**Fig. 1a**). By running IGI-FAST as a research study, we were able to quickly test modifications to our protocol to improve the assay using real-world samples. Pre-mixing saliva samples with 100mM dithiothreitol (DTT) and 10μL of proteinase K (MagMAX Viral/Pathogen Nucleic Acid Isolation kit) prior to nucleic acid extraction improved the rate of RNA extraction success; however a two-hour 50°C pre-incubation and subsequent dilution of saliva samples in DNA/RNA Shield, as presented here, was required to achieve extraction failure rates of <10% (data not shown). We also found that keeping specimens upright at ambient temperature (rather than 4°C) minimized plating failures.

IGI-FAST ran from June through October 2020 and ultimately enrolled a total of 3,653 active participants^7^. Over the course of the study, 11,971 saliva samples were assessed for the presence of SARS-CoV-2 RNA. Participants were invited to provide saliva specimens on alternating weeks at a supervised saliva collection kiosk on campus. During this time, 761 samples (6.4%) were called “Specimen Insufficient”, the majority of which occurred in the first three weeks of the research study when most of the technical protocol optimization as described previously was being performed and research study participants were learning how to provide an appropriate sample (**Fig. 3a**). By comparison, the specimen-insufficient rate of clinically-collected swab specimens processed in our laboratory was relatively stable over the same time period (**Fig. 3b**). Of the 11,210 samples where analysis returned a result, 11,184 were negative for the presence of SARS-CoV-2 RNA, 5 were positive and 21 samples detected only one out of three viral genes and were called inconclusive (**Fig. 3a**). All participants whose saliva returned a positive or inconclusive result were advised to take a clinically-approved test as soon as possible and answer a follow-up survey on the outcome of their clinically-actionable diagnostic result. Of the five participants who received a positive result, two responded to our follow-up survey: one tested positive upon clinical retest and one tested negative. Of the participants who received an inconclusive result, 14 responded to our follow-up survey: 12/14 tested negative upon retest and 2/14 did not seek confirmatory testing (**Supplemental Table 1**). The SARS-CoV-2 ORF1ab and S genes were the most commonly detected viral genes in inconclusive samples, with mean Ct values of 35 (**Supplemental Figure 4**). These high Ct values likely represent non-specific amplification in our RT-qPCR reaction. We have subsequently developed our own in-house RT-qPCR multiplexed primer/probe reagents that provide more accurate viral detection with minimal inconclusive results^11^.

**Figure 3:**
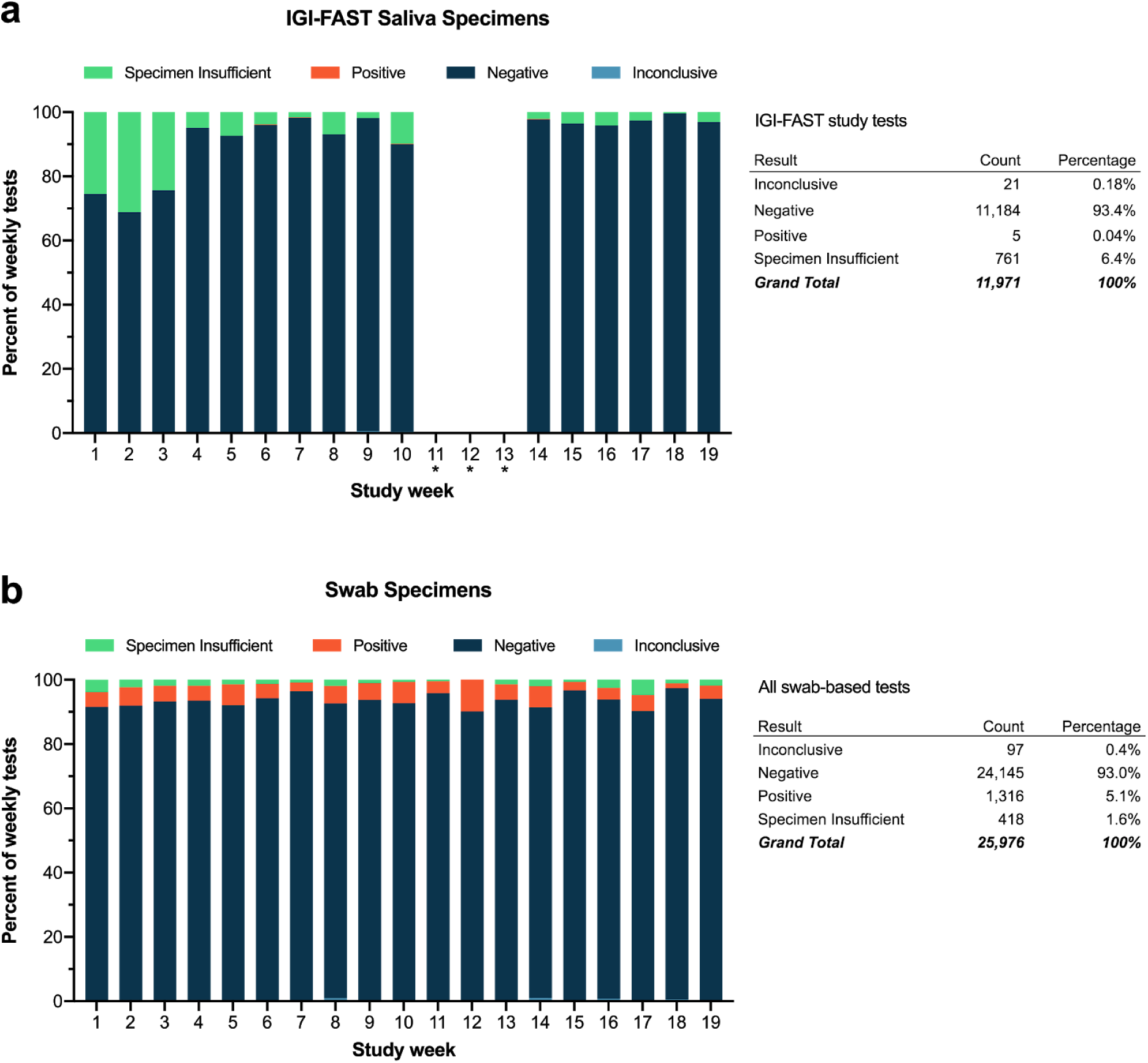
Comparison swab and IGI-FAST saliva specimen results over time. **a**, Final results for saliva specimens collected from asymptomatic individuals through the IGI-FAST study by week. Some samples were collected on week 11 but never processed in the laboratory due to a pipet tip shortage that lasted through weeks 12 and 13. **b**, Final results for all symptomatic swab-based tests run at IGI during the same timeframe as the IGI-FAST study. “∗” indicates weeks IGI-FAST was suspended due to filter pipet tip shortages.

### Four-plex saliva pooling to increase throughput while maintaining test sensitivity

We used samples collected during the IGI-FAST research study to establish a four-plex sample pooling protocol with a goal of increasing sample processing capacity. In our non-pooled protocol, 112µL of a single saliva sample is pipetted by a Hamilton Microlab STARlet liquid handler into a well of a deep-well plate containing 338µL DNA/RNA Shield, and samples that are too viscous to pipette trigger an error warning. Initial experiments to pipette 28µL (1/4 volume of 112µL) from four saliva sample tubes into a single well resulted in pipetting errors that did not trigger a liquid handling warning, leading to pools with unidentifiable missing saliva samples. We therefore moved to a strategy where we generated deep-well extraction plates according to our non-pooled protocol and subsequently combined four plates of saliva-DNA/RNA Shield samples in equal volumes using a Hamilton Microlab VANTAGE 2.0 to generate the final pooled extraction plate. RNA was subsequently extracted from specimens on this pooled extraction plate using the Hamilton Microlab VANTAGE. The saliva sample pooling protocol is described in **Supplemental Figure 5**.

To confirm that *bona fide* positive saliva samples could be identified in four-plex pools, we generated contrived pools with specific combinations of known positive and negative saliva samples previously tested for SARS-CoV-2 RNA in our standard assay. We tested 8 pools consisting of a single positive sample combined with 3 negative samples, as well as 81 pools of only negative samples. Because the goal of a pooled test is to flag pools that may contain a positive sample, we reasoned that an inconclusive or positive pool result would be sufficient to flag a pool for retesting. Following extraction and RT-qPCR, all 8 pools were flagged (7 positive and 1 inconclusive), yielding a positive agreement of 100% (**Fig. 4a**). When comparing Ct values for the positive pools and the positive samples analyzed individually, we observed a consistent increase upon pooling as expected (**Fig. 4b**). This reduced sensitivity upon pooling could be due to sample dilution and/or to degradation of the sample between the original test and pooling.

**Figure 4:**
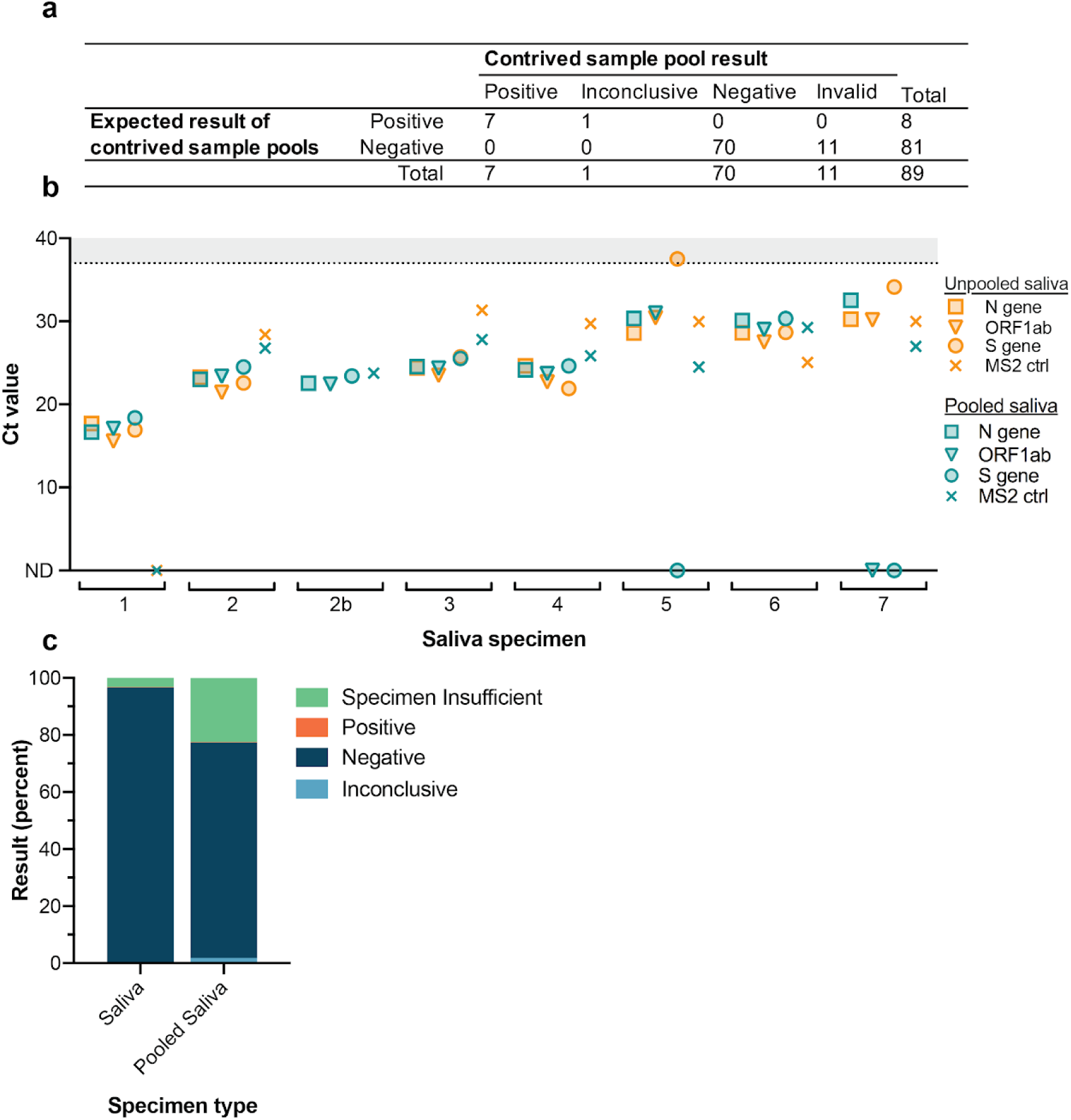
Four-plex pooling of saliva specimens. Four-plex pools were generated with saliva samples previously determined to be positive or negative for the presence of SARS-CoV-2 RNA. 81 wells were generated to contain four negative samples and eight contained one positive sample mixed with 3 negative samples. **a**, All wells containing positive sample pools were called either positive (7/8) or inconclusive (1/8). **b**, Viral and MS2 Ct values for unpooled and pooled positive saliva samples. Note that unpooled saliva sample 2 was run in two separate positive pools while all other positive pools contained unique positive samples. Samples were sorted by the N gene Ct value in unpooled saliva samples. Ct values >37 are shaded in gray. Undetected Ct values are plotted as zero and designated by “ND”, not detected. **c**, Results of pooled samples are compared to the final results for unpooled saliva samples run on the finalized saliva extraction protocol. “Specimen Insufficient” pooled samples were subsequently re-run either in new pools or unpooled to obtain a final result.

We ran IGI-FAST participant samples using four-plex pooling during weeks 14-16 of the IGI-FAST study and detected a positive saliva pool immediately upon implementation. We anticipated that, in practice, pooled saliva samples would exhibit a reduced specimen-insufficient rate compared to that of unpooled saliva because analysis of challenging saliva specimens would be facilitated by dilution with other samples in the pool. In contrast to this expectation, upon pooling assay implementation we observed 22.5% of saliva pools were called “Specimen Insufficient”, compared to 3.3% of unpooled saliva samples resulted on the finalized saliva extraction protocol (**Fig. 4c**). All pools called “Specimen Insufficient” had to be re-run, either in new pools or individually, to obtain a final sample result. This high rate of re-running pools (in this case due to specimen insufficiency rather than a high positivity rate) canceled any expected advantage of pooling on laboratory capacity. In addition to lending no added capacity, the high specimen-insufficient rate of saliva pools had the undesirable effect of increasing the turn-around time of sample result determination and was logistically difficult to manage in the laboratory. We subsequently returned to running any saliva specimens as single samples.

### Concordance studies to determine saliva test reproducibility

To determine clinical utility of our saliva testing protocol, we compared virus detection in saliva to that measured in nasopharyngeal (NP) swab samples from the same individuals. Paired saliva and NP specimens were collected through an outpatient tent at Washington Hospital Healthcare System (WHHS) where the hospital provided testing services to symptomatic individuals. Upon consent (**Supplemental Figure 6**), individuals were asked to provide saliva specimens if they had abstained from eating, drinking and smoking for 30 minutes prior to sample collection. To collect paired specimens, the clinician first collected NP swab samples using the hospital’s standard collection kit containing viral transport medium, which would be used to diagnose the patients. Immediately following NP collection, patients were guided through saliva sample collection using the OMNIgene saliva collection method. In order to directly compare saliva and NP as specimen types, we sought to process both specimens in our laboratory using the same extraction and PCR methods. To generate an NP specimen that could be processed at IGI’s clinical laboratory, the WHHS clinical laboratory transferred an aliquot of the NP swab sample into the IGI’s swab sample collection medium (resulting in a 1:1 dilution). The remaining sample was sent to ARUP Laboratories for analysis and clinical diagnosis of the patients (**Fig. 5a**). This process thus produced three test results from each individual: ARUP NP, IGI NP, and saliva. Samples from 128 individuals were collected and 19 individuals were determined to be positive by ARUP NP, IGI NP, and/or saliva (**Fig. 5b**). When comparing positives detected in IGI NP swab and saliva assays, we observed a 73% positive percent agreement (PPA) and a 97% negative percent agreement (NPA) (**Fig. 5c**). Of the 19 positive ARUP tests, 11 were identified as positive by saliva, 2 were flagged as inconclusive, and 1 consistently failed to amplify any genes including the MS2 control. This comparison yielded a 69% PPA and a 100% NPA (**Fig. 5d**).

**Figure 5:**
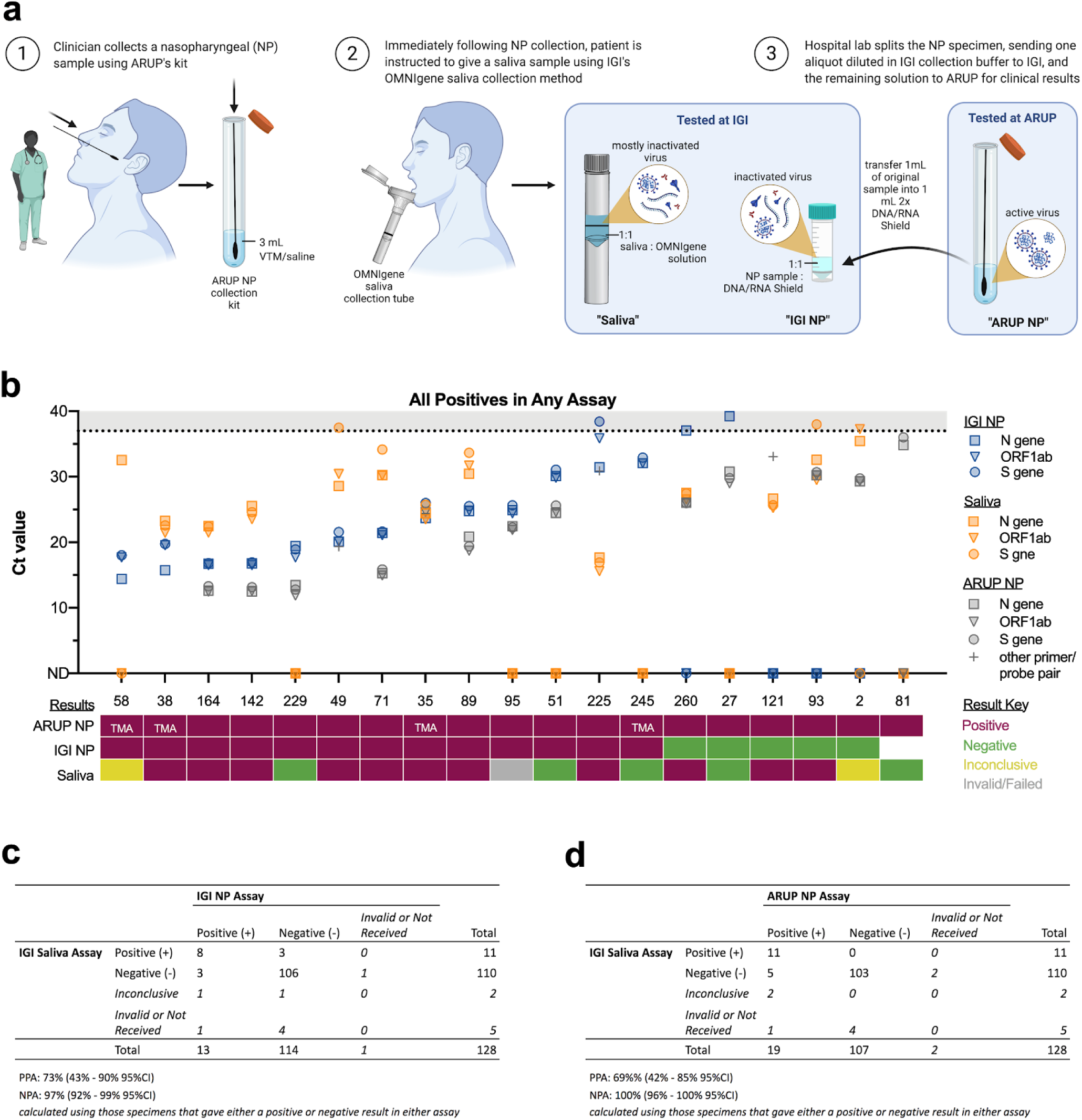
Clinical concordance between IGI-FAST saliva and NP swab samples. The IGI partnered with Washington Hospital Healthcare System (WHHS) to collect paired nasopharyngeal (NP) swab and saliva specimens to assess the concordance between NP swab and saliva-based tests for detection of SARS-CoV-2. **a**, A schematic of how paired samples were collected and split for analysis. Note that undiluted NP samples were analyzed at ARUP Laboratories (ARUP NP) whereas NP samples analyzed at IGI (IGI NP) were diluted 1:1 with DNA/RNA Shield prior to extraction. **b**, Viral Ct values for IGI NP, saliva samples, and ARUP (where available) (top) and the final result of ARUP NP, IGI NP, and saliva samples (bottom). TMA indicates the sample was analyzed using transcription-mediated amplification, thus no Ct values were generated. Samples were sorted by the IGI NP N gene Ct value. As only positive samples are presented, MS2 Ct values were omitted for clarity. Ct values >37 are shaded in gray. Undetected Ct values are plotted as zero and designated by “ND”, not detected. An aliquot of the NP sample for patient 81 was never received by IGI. **c**, Concordance between the IGI saliva and IGI NP and **d**, IGI saliva and ARUP NP assays. Note: some individual samples had no result (invalid or no sample/result received) and were thus left out of Positive Percent Agreement (PPA) and Negative Percent Agreement (NPA) calculations.

Each NP specimen was tested by two different laboratories - the IGI and ARUP Laboratories. NP specimens sent to ARUP were processed by one of at least three different EUA-awarded assays. Of those that returned positive results, ARUP supplied the Ct values when available (**Fig. 5b**). Examining Ct values reveals a strong correlation between the two laboratories for the same NP specimens (**Fig. 5b** and **Supplementary Figure 7a**) with a shift to lower Ct values in ARUP’s results. This shift is partially explained by the necessary dilution of the ARUP NP samples into IGI’s collection buffer in order to process the samples at IGI, but also suggests that ARUP is employing a more sensitive test. Of the 18 ARUP-positive NP specimens that were tested at IGI, 5 returned negative results. These 5 specimens were also those with the highest ARUP Ct values and thus contained low viral concentrations, which once diluted into IGI collection buffer, were likely outside the IGI’s swab assay detection limit.

In contrast, saliva samples that were discordant with their paired NP results were not restricted to those with low NP viral concentrations. Two out of five patients with the highest viral concentrations in their NP samples (Ct values below 20) returned negative or inconclusive results with saliva (**Fig. 5b**, patient 58 and 229 out of patients 58, 38, 164, 42, 229, and 49), while of the eight patients with the lowest NP viral loads (Ct values not detected or above 30 in IGI’s assay and above 25 in ARUP’s assays), half were nonetheless positive in their saliva samples with varying concentrations of virus detected (**Fig. 5b**, patients 225, 260, 121, and 93). We speculate that the observed discordance between saliva and NP results (**Fig. 5c**,**d**) may be attributable to biological factors, such as differences in the location of viral shedding during the stage of disease when samples were collected. This hypothesis is further supported by the lack in overall correlation between Ct values in paired saliva and NP samples (**Supplemental Figure 7b**). Without further data however, we cannot rule out other factors derived from differences in the assay.

## Discussion

The COVID-19 pandemic has required rapid scientific innovation and public health intervention. University campuses have a high density of individuals in an age bracket where SARS-CoV-2 infection generally results in asymptomatic or mild disease, but whose travel and community exposure may increase risk of virus transmission. We previously established a pop-up SARS-CoV-2 clinical laboratory at the University of California, Berkeley to test oropharyngeal swabs collected from symptomatic individuals, both from our campus and surrounding community^6^. Here, we developed an automated pipeline to detect SARS-CoV-2 from saliva samples. Our saliva test demonstrates robust performance with an LoD of 3×10^3^ RNA copies/mL and identified positive cases during four months of use with asymptomatic/presymptomatic study participants.

Our work identified important advantages and disadvantages of SARS-CoV-2 detection from saliva. Robust robotic nucleic acid extraction from saliva enabled concentration of nucleic acids from each sample, allowing sensitive detection and sample pooling. However, we found that the highly variable nature of saliva quality in practice affects extraction and amplification processes. Despite saliva sample pre-treatment, we observed a specimen-insufficient rate higher than that of respiratory swab samples and unacceptably high to run four-plex saliva pooling at scale. The unusual nature of this sample matrix type is underscored by our observation (and that of others through personal communication) that when a saliva specimen is stored for two or more days, it often becomes more tractable, posing minimal problems with sample pipetting and nucleic acid extraction. Operating IGI-FAST as a research study allowed for assay optimization on freshly-collected saliva specimens^7^; we found that this was critical in developing a reliable assay for the robotic isolation of nucleic acid from saliva. A key takeaway from this work is the importance of performing validation experiments with freshly-collected saliva samples from the target population to ensure assay performance is assessed according to real-world applications of the assay.

Although saliva is an easily collected specimen, efforts to optimize the assay were unable to match the clinical sensitivity of our swab-based method. Further, when assessing the clinical sensitivity, we observed discordance between saliva and NP specimens, both in our own laboratory, and with external laboratory results (73% positive percent agreement when comparing IGI NP and saliva assays), which may be explained by several factors. First, the discordance could be biological and attributable to differences in viral loads at these sites in the body or the stage of infection at time of sample collection. In fact, differences have been documented elsewhere when comparing Ct values between NP and saliva specimens^9,12–15^. Further, when comparing our results to other published saliva/NP concordance studies and EUAs, we found that our patient population and collection criteria differed significantly from other publications; some consisted of inpatient samples, were obtained following pre-screening for SARS-CoV-2 by NP swab, or using sputum-laden saliva specimen involving coughing material forward into the mouth^9,14,16,17^. These differences in study design may explain how we observe a lower PPA between saliva and NP than published elsewhere. Of note, we had difficulty consenting inpatients as many of these patients were too infirm to provide a saliva sample, highlighting the fact that active patient participation in specimen collection is required for saliva. Interestingly, while not widely published, a number of laboratories have also experienced marked challenges with saliva as a specimen type and have subsequently determined to continue with swab-based assays as a result^18^. Future studies will directly compare our saliva assay to saliva-based surveillance assays implemented on other university campuses.

In order to expand and unify different testing methods on the UC Berkeley campus, we have moved to a supervised self-swab method that is amenable to sample pooling and has enabled the IGI Clinical Laboratory to continue asymptomatic surveillance efforts at a larger scale. Despite its challenges, saliva remains an important alternative to swab-based assays. We continue to offer the saliva assay described here as a surveillance test, often in a take-home setting, in order to enable testing of smaller target groups who are difficult to recruit into larger surveillance programs or who have difficulties tolerating swab-based collection. For example, a UC Berkeley surveillance project recently identified SARS-CoV-2 in wastewater draining from university-owned housing units^19,20^, which triggered the rapid deployment of saliva-based surveillance testing to residents. In conclusion, we have found detection of SARS-CoV-2 from saliva to be an important alternative sample collection method that complements swab-based diagnostic and surveillance testing.

## Methods

### IGI-FAST saliva sample collection, robotic nucleic acid extraction and RT-qPCR detection of SARS-CoV-2

UC Berkeley students, postdoctoral fellows, staff and faculty working on campus were invited to enroll in the IGI-FAST study (IRB 2020-05-13336). Participants were observed during saliva sample collection at one of two campus kiosks to ensure participants provided the appropriate volume of saliva (1mL) into OMNIgene sample collection tubes (OM-505, DNA Genotek) and correctly deployed the collection tube buffer to mix with the saliva. Upright saliva sample tubes were then heated at 50°C for 120 minutes followed by 65°C for 30 minutes in incubators before being transported into the laboratory.

Detailed protocols for extracting nucleic acids from saliva are presented in Supplemental Figure 3 (unpooled saliva samples) and Supplemental Figure 5 (pooled saliva samples). In brief, 112μL of unpooled saliva samples were added to wells of a deep-well extraction plate (P-DW-11-C-S, Axygen) containing 338μL of 2x DNA/RNA Shield (11-358DB, Zymo) using a Hamilton STARlet liquid handling robot. Up to four deep-well extraction plates were transferred to the Hamilton VANTAGE 2.0 liquid handling robot and the MagMAX Viral/Pathogen Nucleic Acid Isolation Kit (A42352, ThermoFisher), which was previously adapted for use on the Hamilton VANTAGE 2.0^6^, was used for nucleic acid extraction with slight modifications: increased volume of magneticbead-containing binding buffer (20μL beads+530μL binding buffer) and elution solution volume of 30μL. The Hamilton VANTAGE 2.0 was subsequently used to transfer 5μL of eluate to wells of a pre-made and validated 384-well RT-qPCR plate containing 7.5μL TaqPath COVID-19 Combo Kit reagent master mix (A47814, ThermoFisher) to reach a final volume of 12.5μL per well. Sealed RT-qPCR plates were then vortexed for a minimum of 25 seconds before being centrifuged and analyzed on a QuantStudio-6 (ThermoFisher) by RT-qPCR. The TaqPath COVID-19 Combo Kit (ThermoFisher) logic was used for calling samples “positive”, “inconclusive”, “negative”, and “invalid” (see Figure 1a). A “Specimen Insufficient” outcome could arise from: (1) an inappropriate sample volume in the collection tube; (2) a leak in the specimen collection bag; (3) a failure to pipet during sample plating; or (4) a failure to extract RNA.

### Viral inactivation assays

In the BSL-3, 500μL SARS-CoV-2 (3.6-1.58×10^6^ TCID50/mL) was mixed 1:1 with 500μL of OMNIgene buffer and incubated for the indicated amounts of time. Following incubation, the 1mL virus/OMNIgene mix was divided to inoculate eight T175 flasks containing Vero-E6 cells at 80% confluency (125ul/flask). OMNIgene buffer was mixed 1:1 with phosphate buffered saline (PBS) and 125μL was used to inoculate one T175 flask as a negative control and SARS-CoV-2 was mixed 1:1 with PBS and used to inoculate one T175 flask with 125μL as a positive control. On day 3 post inoculation, cytopathic effect (CPE) was assessed and 5mL of supernatant from each flask was transferred into a new set of Vero-E6 flasks at 80% confluency. CPE was again assessed at day 7 post initial inoculation (4 days post inoculation of the second set of flasks).

### Optimization of DNA/RNA Shield addition to maximize nucleic acid extraction

For optimizing extraction of contrived positive samples (Fig. 1c), TaqPath COVID-19 Control RNA (ThermoFisher, A48003) was spiked into saliva collected from four individual donors at 5×10^3^copies/ml, then 1mL was added to OMNIgene OM-505 collection tubes (DNA Genotek) and mixed 1:1 with the OMNIgene collection tube buffer. Sample tubes were inverted 10 times to mix and then 450μL of each sample arrayed by hand into a deep well extraction plate (for undiluted samples) or 270μL 2x DNA/RNA Shield was mixed with 180μL saliva/RNA (for 3:2 diluted samples). Contrived samples were subjected to nucleic acid extraction and RT-qPCR analysis as described and MS2 and viral genes were used to assay sample extraction efficiency. For optimizing the extraction of saliva samples previously reported “insufficient” in the IGI-FAST study (Supplemental Fig. 2), samples were diluted in either PBS or 2x DNA/RNA Shield and the RT-qPCR detection of spiked-in MS2 was used to quantify sample extraction efficiency.

### Saliva assay validation assays

Saliva was collected in OM-505 collection tubes (DNA Genotek) from four unique donors negative for SARS-CoV-2 by swab. These samples were used to generate titration curves with TaqPath COVID-19 Control RNA (ThermoFisher, A48003) or heat-inactivated virus (Stanley laboratory, UC Berkeley) to determine the assay’s limit of detection (LoD). To test the robustness and reproducibility of our assay, the LoD was confirmed by generating unique saliva samples (previously determined to be negative for SARS-CoV-2) at 1x, 2x and 5x the SARS-CoV-2 LoD.

### Saliva pooling

The saliva four-plex pooling protocol was validated by creating contrived pools with saliva samples known to be positive or negative for the presence of SARS-CoV-2. Eighty-one wells were generated to contain four negative samples per well (324 negative samples) and eight wells were generated to contain one positive sample mixed with three negative samples (8 positive samples + 24 negative samples). Note that two contrived positive pools contained the same positive sample, while the remaining six contrived positive wells contained unique positives.

Saliva sample pooling was implemented in weeks 14, 15, and 16 of the IGI-FAST research study. The “Specimen Insufficient” rates of pooled and unpooled saliva samples was calculated for all samples run on the finalized saliva extraction protocol (which included the 120 minute 50°C pre-incubation and DNA/RNA Shield sample dilution). The “Specimen Insufficient” rate was calculated for all pooled samples in weeks 14-16, and for all unpooled saliva samples in weeks 8-10 and 17-19. The pooled sample results may differ from their final sample result, as pooled samples that returned a “specimen insufficient” result were re-run in new pools or as unpooled samples until a final result was obtained.

### Washington Hospital Healthcare System (WHHS) nasopharyngeal swab/saliva concordance experiment

Through a collaboration with WHHS, we obtained paired nasopharyngeal (NP) swab and saliva specimens from symptomatic individuals. These individuals consented at a walk-up testing site and were accepted if the patient had abstained from eating, drinking and smoking for 30 minutes prior to sample collection. The WHHS institutional review board (IRB) waiver and consent form for NP/saliva comparison are presented in **Supplemental Figure 6**. To collect paired specimens, the clinician first collected an NP swab sample into viral transport medium, followed immediately by saliva sample collection using our established OMNIgene saliva collection method described above. Within 2 hours, personnel at the WHHS clinical laboratory transferred 1mL of swab-derived sample medium into the IGI’s swab collection tube containing 1mL 2x DNA/RNA Shield. The remaining sample medium with the swab was sent to ARUP Laboratories for analysis. Undiluted NP samples were analyzed at ARUP Laboratories (ARUP NP) on one of their EUA-holding SARS-CoV-2 (COVID-19) assays whereas NP samples analyzed at IGI (IGI NP) were diluted 1:1 with 2x DNA/RNA Shield prior to extraction, then analyzed according to our previously published swab-based assay^6^.

## Data Availability

The datasets generated for the current study are available upon reasonable request.

## Acknowledgements

Thank you to Xammy Huu Nguyenla and Erik Van Dis of Sarah Stanley’s laboratory at the University of California, Berkeley for generating the heat-inactivated virus and performing the viral inactivation studies. Thank you to the additional members of IGI SARS-CoV-2 Testing consortium: Alexandra M. Amen, Kerrie W. Barry, John M. Boyle, Cara E. Brook, Seunga Choo, L.T. Cornmesser, David J. Dilworth, Indro Fedrigo, Skyler E. Friedline, Thomas G. W. Graham, Ralph Green, Megan L. Hochstrasser, Dirk Hockemeyer, Netravathi Krishnappa, Azra Lari, Hanqin Li, Tianlin Lu, Elijah F. Lyons, Kevin G. Mark, Lisa Argento Martell, A. Raquel O. Martins, Patrick S. Mitchell, Christine Naca, Divya Nandakumar, Elizabeth O’Brien, Derek J. Pappas, Diana L. Quach, Benjamin E. Rubin, Rohan Sachdeva, Abdullah Muhammad Syed, I-Li Tan, Amy L. Tollner, Timothy K. Turkalo, M. Bryan Warf, and Oscar N. Whitney. Thank you to the Washington Hospital Healthcare System Emergency Department Rapid Screening and Treatment Unit (RSTU) and Clinical Laboratory for assistance with saliva sample collection and processing.

