## Supplemental Figures for "Robotic RNA extraction for SARS-CoV-2 surveillance using saliva samples"

| SARS-CoV-2 | OMNIgene | Incubation | CPE, 3 dpt | CPE, 7 dpt |
|---|---|---|---|---|
| + | + | 120 min 50°C | 0/8 CPE+ | 0/8 CPE+ |
| + | + | 120 min 56°C | 0/8 CPE+ | 0/8 CPE+ |
| + | - | - | 1/1 CPE+ | 1/1 CPE+ |
| - | + | - | 0/1 CPE+ | 0/1 CPE+ |

**Supplemental Figure 1: Heat inactivation of SARS-CoV-2 in OMNIgene solution —** Cultured SARS-CoV-2 ($3.16x10^6$ TCID50/ml) was mixed 1:1 with OMNIgene solution present in OM-505 collection tubes to test incubation conditions that inactivate viral replication. Samples were either incubated at 50°C or 56°C for the indicated length of time before being applied to Vero-E6 cells. Cytopathic effect (CPE) was quantified at 3 and 7 days post treatment (dpt).

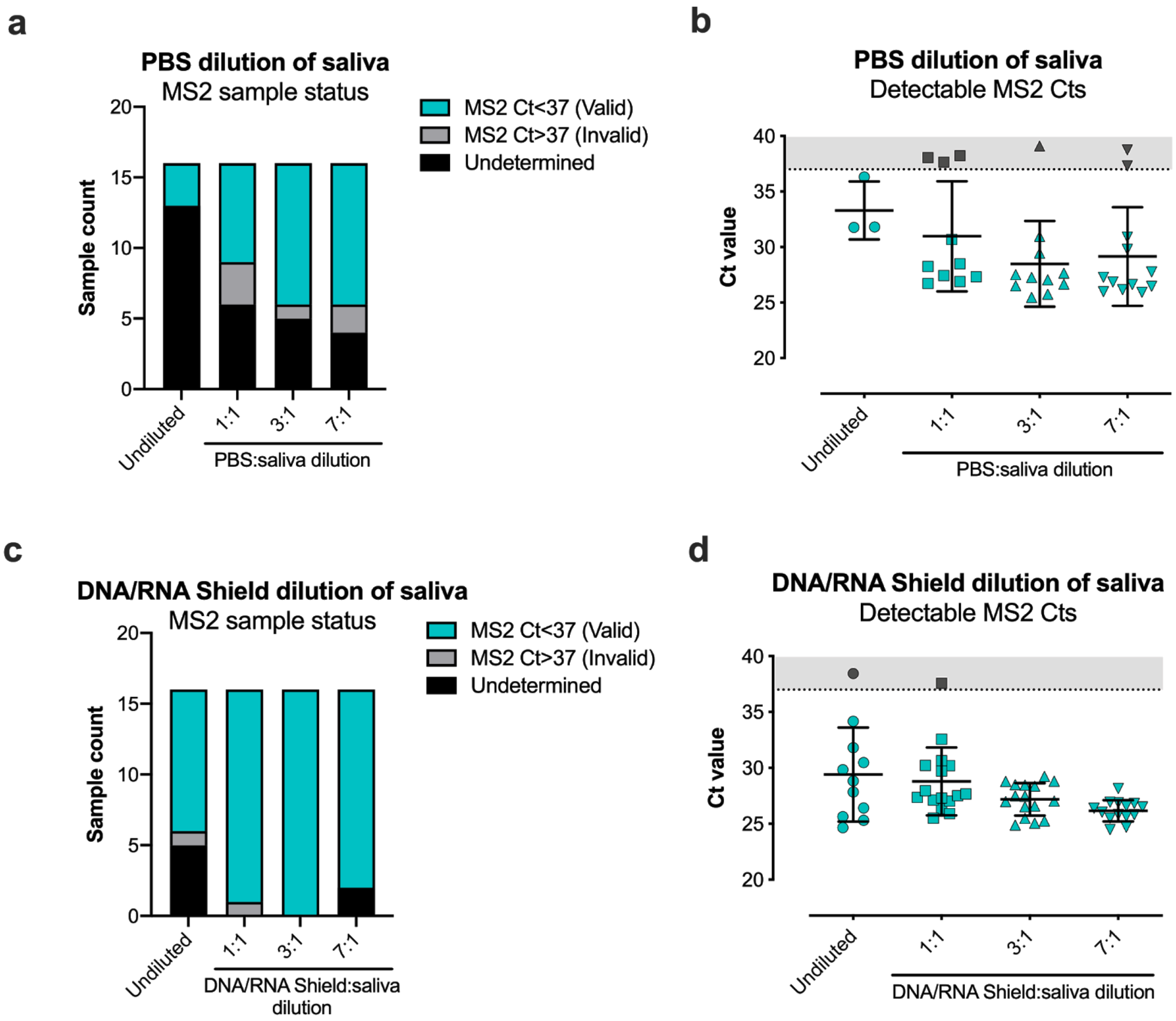


**Supplemental Figure 2: Optimizing the dilution of saliva samples to maximize RNA extraction —** Saliva samples previously reported “specimen insufficient” in the IGI-FAST study were diluted in either phosphate buffered saline (PBS) or 2x DNA/RNA Shield and the RT-qPCR detection of spiked-in MS2 was used to quantify sample extraction efficiency. **a**, Sample status after qRT-pCR for MS2 after serial dilution in PBS. The same set of 16 saliva samples were used for each dilution condition. **b**, MS2 Ct values for samples diluted in PBS. **c**, Sample status after qRT-pCR for MS2 after serial dilution in DNA/RNA Shield. The same set of 16 saliva samples were used for each dilution condition (distinct from the samples used for **a**). **d**, MS2 Ct values for samples diluted in DNA/RNA Shield. For **b** and **d,** the mean and standard deviation are plotted for each group and invalid MS2 Ct>37 are indicated in gray.

**Supplemental Figure 3: Protocol for the robotic nucleic acid extraction from saliva and RT-qPCR detection of SARS-CoV-2 RNA**

**Sample collection and pre-extraction processing**

1. 1mL saliva collected in DNA Genotek OMNIgene OM-505 sample tube
2. Snap cap shut to release 1mL Genotek Solution and invert by hand 10 times
3. Heat tubes at 50°C for 2 hours in collection tube
4. Heat inactivate at 65°C for 30 min in collection tube

**Hamilton Starlet sample accessioning**

1. Starlet pipettes 338μL 2x Zymo DNA/RNA Shield into a 96-well deep-well extraction plate
2. Starlet pipettes 112μL saliva sample or extraction control into respective wells
    a. Extraction controls
        i. Negative control: PBS mixed 1:1 with 2x DNA/RNA Shield
        ii. Human matrix control: 200ng/ml human RNA in PBS mixed 1:1 with 2x DNA/RNA Shield
        iii. Positive control: Thermo Fisher TaqPath COVID-19 Control (Cat # A48003) diluted 1:1000 in PBS mixed 1:1 with 2x DNA/RNA Shield

**Hamilton Vantage Extraction** using the MagMAX Viral/Pathogen Nucleic Acid Isolation Kit (ThermoFisher, A42352)

***Bead binding***

1. 96-channel core head adds 10μL MS2 to each well
2. 96-channel core head adds 10μL Proteinase K to each well
3. 96-channel core head adds 550μL binding buffer + bead mixture to each well
    a. Per well:
        i. 20μL beads
        ii. 530μL binding buffer
4. 96-channel core head mixes reagents
5. Extraction plate sits for 5 minutes at room temperature
6. Beads in the extraction plate are collected on a Alpaqua Magnum FLX magnet for 10 minutes
7. 96-channel core head slowly aspirates supernatant

***Bead washing***

*Wash 1*

8. 96-channel core head adds 750μL wash buffer and mixes
9. Beads in the extraction plate are collected on a Alpaqua Magnum FLX magnet for 5 minutes
10. 96-channel core head slowly aspirates supernatant

*Wash 2*

11. Plates are transferred off the magnet
12. 96-channel core head adds 750μL 80% ethanol and mixes
13. Beads in the extraction plate are collected on a Alpaqua Magnum FLX magnet for 5 min
14. 96-channel core head slowly aspirates supernatant

*Wash 3*

15. Plates are transferred off the magnet.
16. 96-channel core head adds 500μL 80% ethanol and mixes
17. Beads in the extraction plate are collected on a Alpaqua Magnum FLX magnet for 2 min

18. 96-channel core head slowly aspirates supernatant

***Elution***

19. Extraction plate is moved to HSS and shakes for 2 minutes (to dry beads)
20. 96-channel core head adds 30µL elution buffer
21. Extraction plate is moved to HSS and shakes for 2 minutes
22. Extraction plate is incubated at 65C for 10 minutes (off deck)
23. Extraction plate is moved to HSS and shakes for 5 minutes
24. Beads in the extraction plate are collected on a Alpaqua Magnum FLX magnet for 2 minutes
25. 96-channel core head transfers 22µL of elution from the extraction plate to an elution plate

**Hamilton Vantage RT-qPCR plate setup**

1. 5µL of eluate or control (nuclease-free water or Thermo FIsher COVID19 positive RNA) is distributed to previously made and validated 384-well RT-qPCR plates containing TaqPath COVID-19 Combo Kit reagent (A47814)
    a. Per well:
        i. 3.125µL TaqPath 1-Step Multiplex Master Mix (No ROX) 4x
        ii. 0.625µL COVID-19 Real Time PCR Assay Multiplex Primers and Probes
        iii. 3.75µL Sterile Nuclease-Free Water
2. Plate is sealed, vortexed for a minimum of 25 seconds, spun down and run on a QuantStudio-6

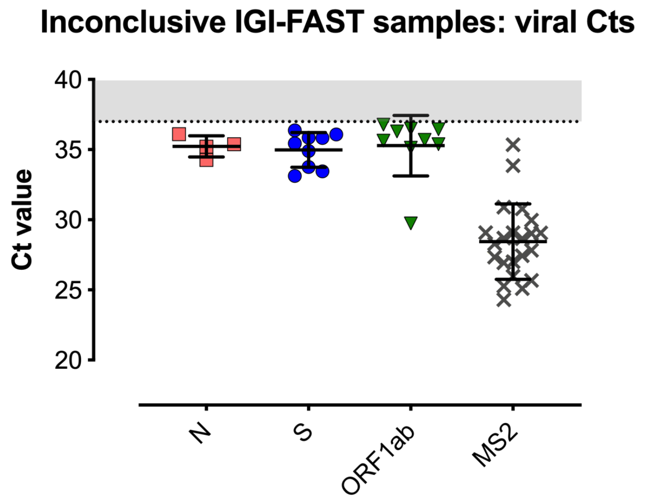


**Supplemental Figure 4: Viral genes detected in inconclusive samples —** Viral and MS2 Cts for IGI-FAST saliva samples with an inconclusive result (one viral gene and MS2 detected at a Ct value <37). Mean and standard deviation are plotted for each group. Gray shading indicates Ct values >37.

**Supplemental Figure 5: Four-plex pooling of saliva samples for robotic nucleic acid extraction and RT-qPCR detection of SARS-CoV-2 RNA**

*Note: Differences between singlet and four-plex pooling assays are highlighted*

**Sample collection and pre-extraction processing**

1. 1mL saliva collected in DNA Genotek OMNIgene OM-505 sample tube
2. Snap cap shut to release 1mL Genotek Solution and invert by hand 10 times
3. Heat tubes at 50°C for 2 hours in collection tube
4. Heat inactivate at 65°C for 30 min in collection tube

**Hamilton Starlet sample accessioning**

3. Starlet pipettes 338μL 2x Zymo DNA/RNA Shield into a 96-well deep-well extraction plate
4. Starlet pipettes 112μL saliva sample or extraction control into respective wells
    a. Extraction controls
        i. Negative control: PBS mixed 1:1 with 2x DNA/RNA Shield
        ii. Human matrix control: 200ng/ml human RNA in PBS mixed 1:1 with 2x DNA/RNA Shield
        iii. Positive control: Thermo Fisher TaqPath COVID-19 Control (Cat # A48003) diluted 1:1000 in PBS mixed 1:1 with 2x DNA/RNA Shield

**Hamilton Vantage four-plex mixing of extraction plates**

5. Four extraction plates containing saliva + DNA/RNA Shield are loaded onto the Hamilton Vantage
6. An empty extraction plate is loaded onto the Hamilton Vantage
7. 96-channel core head gently mixes and moves 112μL from all wells of each extraction plate into the empty extraction plate to pool four samples per well

**Hamilton Vantage Extraction** using the MagMAX Viral/Pathogen Nucleic Acid Isolation Kit (ThermoFisher, A42352)

***Bead binding***

26. 96-channel core head adds 10μL MS2 to each well
27. 96-channel core head adds 10μL Proteinase K to each well
28. 96-channel core head adds 550μL binding buffer + bead mixture to each well
    a. Per well:
        i. 20μL beads
        ii. 530μL binding buffer
29. 96-channel core head mixes reagents
30. Extraction plate sits for 5 minutes at room temperature
31. Beads in the extraction plate are collected on a Alpaqua Magnum FLX magnet for 10 minutes
32. 96-channel core head slowly aspirates supernatant

***Bead washing***

*Wash 1*

33. 96-channel core head adds 750μL wash buffer and mixes
34. Beads in the extraction plate are collected on a Alpaqua Magnum FLX magnet for 5 minutes
35. 96-channel core head slowly aspirates supernatant

*Wash 2*

36. Plates are transferred off the magnet
37. 96-channel core head adds 750µL 80% ethanol and mixes
38. Beads in the extraction plate are collected on a Alpaqua Magnum FLX magnet for 5 min
39. 96-channel core head slowly aspirates supernatant

*Wash 3*

40. Plates are transferred off the magnet.
41. 96-channel core head adds 500µL 80% ethanol and mixes
42. Beads in the extraction plate are collected on a Alpaqua Magnum FLX magnet for 2 min
43. 96-channel core head slowly aspirates supernatant

***Elution***

44. Extraction plate is moved to HSS and shakes for 2 minutes (to dry beads)
45. 96-channel core head adds 30µL elution buffer
46. Extraction plate is moved to HSS and shakes for 2 minutes
47. Extraction plate is incubated at 65C for 10 minutes (off deck)
48. Extraction plate is moved to HSS and shakes for 5 minutes
49. Beads in the extraction plate are collected on a Alpaqua Magnum FLX magnet for 2 minutes
50. 96-channel core head transfers 22µL of elution from the extraction plate to an elution plate

**Hamilton Vantage RT-qPCR plate setup**

3. 5µL of eluate or control (nuclease-free water or Thermo FIsher COVID19 positive RNA) is distributed to previously made and validated 384-well RT-qPCR plates containing TaqPath COVID-19 Combo Kit reagent (A47814)
    a. Per well:
        i. 3.125µL TaqPath 1-Step Multiplex Master Mix (No ROX) 4x
        ii. 0.625µL COVID-19 Real Time PCR Assay Multiplex Primers and Probes
        iii. 3.75µL Sterile Nuclease-Free Water
4. Plate is sealed, vortexed for a minimum of 25 seconds, spun down and run on a QuantStudio-6

**Supplemental Figure 6: WHHS institutional review board (IRB) waiver and consent form for NP/saliva comparison**

Washington Hospital Healthcare System

2000 Mowry Avenue, Fremont, California 94538-1716 | 510.797.1111
www.whhs.com

To whom it may concern,

As part of our ongoing effort to increase COVID-19 testing and simplify COVID-19 testing for our community we have reviewed a saliva to nasopharyngeal (NP) swab comparison study involving the University of California Berkeley (UCB) Institute of Genomic Innovations (IGI) and for our purposes this study is not considered research by Washington Hospital Healthcare System (WHHS) IRB. As such, a consent form that has been approved by WHHS compliance and the appropriate WHHS legal representative will be sufficient to send de-identified samples to the IGI.

Any concerns can be brought directly to laboratory at WHHS.

Thank you,

Garth Huberty
Director of Laboratory Services
Washington Hospital Healthcare System

office (510) 818-6268 lab (510) 818-3440

*Kimberly Hartz, Chief Executive Officer*
Washington Township Health Care District • Washington Hospital • Institute for Joint Restoration and Research • Sandy Amos RN Infusion Center
Taylor McAdam Bell Neuroscience Institute • UCSF - Washington Cancer Center • Washington Center for Wound Healing & Hyperbaric Medicine
Washington Maternal Child Education • Washington on Wheels • Washington Outpatient Diabetes Program • Washington Outpatient Imaging Center
Washington Outpatient Rehabilitation Center • Washington Outpatient Surgery Center • Washington Prenatal Diagnostic Center
Washington Radiation Oncology Center • Washington Special Care Nursery • Washington Sports Medicine • Washington Township Medical Foundation
Washington Urgent Care • Washington Wellness Center • Washington Women's Center

Washington Hospital Healthcare System

2000 Mowry Avenue, Fremont, California 94538-1716 | 510.797.1111
www.whhs.com

It is our goal at Washington Hospital to provide accurate and timely lab results to all of our patients. In order to deliver on this goal, we often use cutting edge technology, new instruments, and/or new processes. As part of our ongoing professional accreditation we are required to validate any of these changes physically here at the hospital.

Today we are asking for your help in volunteering for a recollection of a nasopharyngeal swab and a saliva sample to compare the two sample collection methods through a collaborative effort with The University of California at Berkeley's Innovative Genomics Institute (IGI).

All identifying information will be removed before sending your samples to the IGI for processing, and your samples will only be used to determine if saliva is appropriate for detecting COVID-19.

It is our mission to improve COVID-19 testing for our patients and the public. There will be no additional charges to you or your insurance.

Your help is greatly appreciated!

I ______________________________ (Print Your Name) agree for Washington Hospital Laboratory to collect additional samples as described above.

Signature: ________________________Date:__________

*Kimberly Hartz, Chief Executive Officer*
Washington Township Health Care District • Washington Hospital • Institute for Joint Restoration and Research • Sandy Amos RN Infusion Center
Taylor McAdam Bell Neuroscience Institute • UCSF - Washington Cancer Center • Washington Center for Wound Healing & Hyperbaric Medicine
Washington Maternal Child Education • Washington on Wheels • Washington Outpatient Diabetes Program • Washington Outpatient Imaging Center
Washington Outpatient Rehabilitation Center • Washington Outpatient Surgery Center • Washington Prenatal Diagnostic Center
Washington Radiation Oncology Center • Washington Special Care Nursery • Washington Sports Medicine • Washington Township Medical Foundation
Washington Urgent Care • Washington Wellness Center • Washington Women's Center

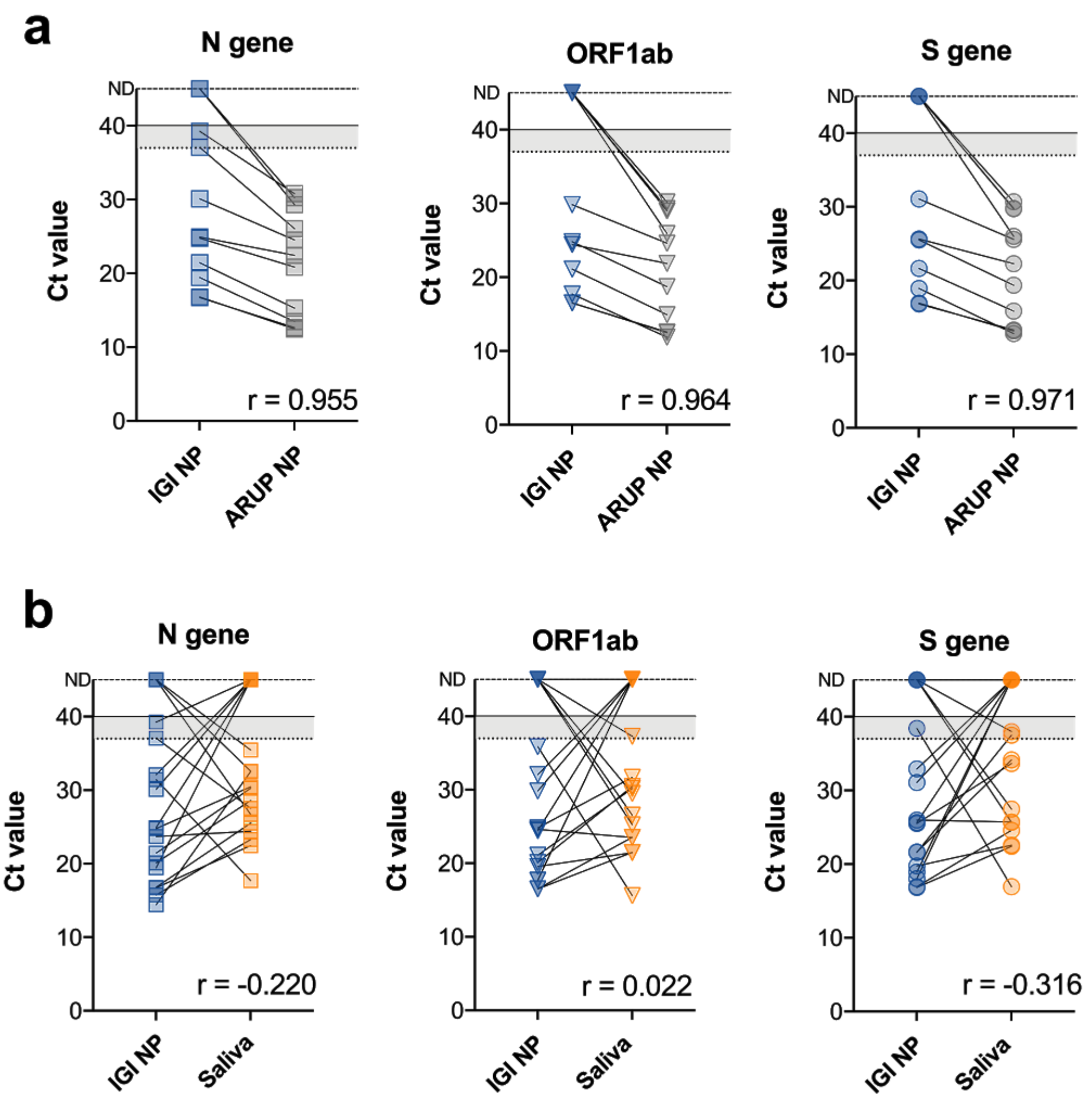


**Supplemental Figure 7 — Correlation between Ct values. a,** Ct values for NP specimens tested by ARUP and IGI using the same primer-probe pairs are plotted with paired results connected by a line. **b**, Ct values for paired NP and Saliva specimens processed by IGI are connected by a line showing only those with paired results in the two assays. Ct values above the 37 cutoff (dotted line) are shaded in gray. PCR was run for 40 cycles. Undetected Ct values are plotted at the top of each graph and designated by “ND”, not detected. Pearson correlation (r) was calculated for each gene between the two indicated assays where undetected Ct values were left blank (no value assigned).
